# Depression, Brain Structure and Socioeconomic Status: A UK Biobank Study

**DOI:** 10.1101/2024.03.27.24304960

**Authors:** Sasha Johns, Caroline Lea-Carnall, Nick Shryane, Asri Maharani

## Abstract

**Background:** Depression results from interactions between biological, social, and psychological factors. Literature shows that depression is associated with abnormal brain structure, and that socioeconomic status (SES) is associated with depression and brain structure. However, limited research considers the interaction between each of these factors.

**Methods:** Multivariate regression analysis was conducted using UK Biobank data on 39,995 participants to examine the relationship between depression and brain volume in 23 cortical regions for the whole sample and then separated by sex. It then examined whether SES affected this relationship.

**Results:** Eight out of 23 brain areas had significant negative associations with depression in the whole population. However, these relationships were diminished in seven areas when SES was included in the analysis. For females, three regions had significant negative associations with depression when SES was not included, but only one when it was. For males, lower volume in six regions was significantly associated with higher depression without SES, but this relationship was abolished in four regions when SES was included. The precentral gyrus was robustly associated with depression across all analyses.

**Limitations:** Participants with conditions that could affect the brain were not excluded. UK Biobank is not representative of the general population which may limit generalisability. SES was made up of education and income which were not considered separately.

**Conclusions:** SES affects the relationship between depression and cortical brain volume. Health practitioners and researchers should consider this when working with imaging data in these populations.

## Introduction

Mental health problems are significant public health issues within the UK and worldwide, with depression being one of the most prevalent. Research shows that there is a relationship between depression and brain structure (Tang et al., 2007; Kronmüller et al., 2009; Bos et al., 2018; Maggioni et al., 2019), and that social factors, including socioeconomic status (SES), are associated with both depression (Delgadillo et al. 2016; Marmot, 2010; Freeman et al., 2016) and changes in brain structure (Kweon et al., 2022; Colich et al., 2020; Jednoróg et al., 2012; Noble et al., 2015). However, these three factors are rarely all considered together. Given the impact of depression on both the individual and society, as discussed below, understanding the mechanisms of this disorder, to improve prevention and treatment options, is of critical importance.

Around 1 in 6 adults in England met the criteria for a common mental health disorder in the week prior to being surveyed (McManus et al., 2016). Mental health problems are one of the leading contributors to overall worldwide disease burden and are reported to account for 16% of Disability Adjusted Life Years (DALYs, Arias et al., 2022). They confer substantial societal and economic implications with an estimated annual cost in England of £77 billion (Kirkwood et al., 2010). Depression is one of the most prevalent mental health problems, affecting approximately 5% of adults (World Health Organization, WHO, 2023) and 280 million people globally (Institute of Health Metrics and Evaluation, 2023). Depression can be long-lasting with a single chronic episode, or recurrent, with many episodes over a long time, and has a substantial impact on an individual’s ability to function and cope with daily life. The risk of suicide is reported to be about 15 times higher in those with depression than the general population (Cipriani et al., 2005). This is however likely to be an underestimate as many who die by suicide have undiagnosed depressive symptoms.

It is widely accepted that mental health problems, including depression, are complex issues caused by biological, social, and psychological factors (Engel, 1997; Kinderman, 2005; Kinderman et al., 2013; Fava & Sonino, 2008). Existing research shows that depression is associated with abnormal brain structure in several brain regions, including reduced volumes of the amygdala and hippocampus, frontal lobe cortical thinning, orbitofrontal thinning, and reduced volume of the precentral gyrus (Tang et al., 2007; Kronmüller et al., 2009; Bos et al., 2018; Maggioni et al., 2019).

Social factors have been shown to be strongly related to mental health problems. SES specifically, has been determined to be both a cause and effect of mental health problems. SES refers to a variety of measures of one’s social class or standing, but most commonly is made up of one’s education, social class, and/or income (Darin-Mattson et al., 2017). Children and adults living in the lowest 20% income bracket in England are 2-3 times more likely to develop mental health problems than those in the highest 20% (Marmot, 2010). Individuals with mental health problems are less likely to excel in school (Schulte-Körne, 2016), and more likely to be unemployed (Egan et al., 2016). In terms of directionality, there is evidence that depression in childhood and adolescence can lead to socioeconomic issues later in life. Using data from the National Longitudinal Study of Youth 1997, Egan et al. (2016) demonstrated that adolescents experiencing high levels of distress were significantly more likely to experience unemployment in later life. There is also evidence for the opposite direction. Lee et al. (2019) conducted a longitudinal study to investigate whether duration of unemployment in young adulthood is associated with mental health problems later and found that duration of unemployment increased mental health problems, whilst accounting for childhood mental and behavioural health problems.

There is also some evidence that social factors are associated with differences in brain structure (Kweon et al., 2022; Colich et al., 2020; Jenoróg et al., 2012; Noble et al., 2015). For example, Kweon et al. (2022) demonstrated that higher SES is related to larger overall grey matter volume (GMV) across the brain, with the strongest regional relationship seen in the cerebellum. Deprivation and SES are associated with thinning of the frontoparietal, default, and visual networks (Colich et al., 2020).

Despite the acknowledgement of a biopsychosocial model of depression, there is limited literature that considers these factors all together. Social factors are not often considered in the context of biological factors. There is a wealth of research highlighting that depression is a major concern in modern society, yet there is still much that is lacking in our understanding of this condition. Given the significant impact of depression on the individual and society, improving understanding of the mechanisms of this disorder is essential to optimise detection, treatment and ultimately prevention.

As discussed, there is an established relationship between brain structure and depression, and this is supported by Harris et al.’s (2022) study using UK Biobank data. They found that different lifetime depression phenotypes were associated with different structural brain regions. The phenotypes they examined were: self-reported depression, whether participants reported a current or past diagnosis of depression; an approximate measure of lifetime depression previously created by Smith et al. (2013), where responses are combined into measures of ‘probable single episode’, ‘probable mild recurrent’ and ‘probable severe recurrent’ (see Smith et al., 2013 for further explanation); and the short form of the Composite International Diagnosis Interview (CIDI-SF), which is based on DSM-IV depression criteria (Diagnostic and Statistical Manual of Mental Health Disorders, fourth edition). They examined associations between brain structural measures and each depression phenotype and found that depression was generally associated with reduced cortical thickness, with self-reported depression particularly showing consistent robust associations with thinner cortex in most regions they examined. They found that regional cortical surface area and volume did not show many significant associations after correcting for multiple comparisons, but most nominally significant results suggested depression was associated with a trend towards greater cortical surface area. Numerous depression phenotypes were associated with reduced grey matter volume in the brain stem and ventral diencephalon, but also with greater volume of the caudate nucleus and putamen. They found fewer significant associations than expected, given the large sample size. They suggested this could be due to focusing on lifetime rather than current major depressive disorder (MDD) (Harris et al., 2022), which may imply that brain morphology changes are expected to be transient and diminish after the MDD episode has resolved. This is in line with research that suggests that brain structure and function may change in response to treatment for or remission of depression (Phillips et al., 2015; Enneking et al., 2020; Kong et al., 2014; Lan et al., 2016; Fu et al., 2004).

This study, therefore, has two main aims; first, to further examine the association between depression measures and brain structure in the UK Biobank. Second, we extended our analysis to test whether the well-established relationship between depression and brain structure is affected by socioeconomic factors such as education and income.

This study extended the existing literature in several ways. Firstly, we used a validated measure of recent depressive symptoms taken at the same time as the brain scans (Recent Depressive Symptoms, RDS-4, Dutt et al., 2022). This allowed us to determine whether the RDS-4 is related to brain structure in the same way as the self-report measures used in Harris et al. (2022), and if more significant associations can be found when using a measure of recent, rather than lifetime, depression. Secondly, we focus on cortical volume in the first instance as this has been shown to be associated with depression in several studies (Belleau et al., 2019; Nolan et al., 2002; Lai et al., 2000; Bremner et al., 2002). Finally, we include education and income in the analysis to examine how this affects the relationship between depression and cortical volume, given that SES is associated with cortical volume (McDermott et al., 2019; Noble et al., 2012; Hair et al., 2015) and depression (Egan et al., 2016; Freeman et al., 2016; Zimmerman & Katon, 2005; Wang et al., 2010).

## Methods

The UK Biobank is a large-scale biomedical database with rich sociodemographic, socioeconomic, mental health and neuroimaging data. It includes over 500,000 participants with an age range of 40-69 years, recruited between 2006-1010. The UK Biobank is particularly useful for this research as it is unique in the fact that it has cross-sectional neuroimaging scans on over 60,000 participants. Our study included a subset of 39,955 UK Biobank participants, who had both structural imaging and recent depression data (RDS-4). Participants who had chosen to withdraw from the UK Biobank study were excluded. See Table 1 for descriptive statistics.

**Table 1:** Descriptive statistics for UK Biobank participants included in analyses.

| Variable | Full sample<br>(n=39,955) | Male<br>(n= 20,930) | Female<br>(n= 19,025) |
| --- | --- | --- | --- |
| Mean Age (SD) | 54.89(7.52) | 55.65(7.62) | 54.20(7.36) |
| Caucasian, % | 97.16% | 97.08% | 97.23% |
| Education category |  |  |  |
| Not higher education, % | 52.33% | 49.60% | 54.09% |
| College or higher, % | 47.67% | 50.40% | 45.91% |
| Income category, %: |  |  |  |
| Less than 18,000 | 11.19% | 9.35% | 12.95% |
| 18,000 to 30,999 | 21.60% | 19.89% | 23.23% |
| 31,000 to 51,999 | 30.32% | 30.68% | 29.97% |
| 52,000 to 100,000 | 28.79% | 31.06% | 26.62% |
| Greater than 100,000 | 8.10% | 9.02% | 7.23% |
| Mean Depression score<br>(SD) | 5.20(1.75) | 5.06(1.62) | 5.34(1.85) |

Brain Magnetic Resonance Imaging (MRI) data were acquired from three dedicated imaging centres with identical 3T Siemens Skyra scanner and 32-channel head coil. The full protocol has been described by Miller et al. (2016). This study used T1-weighted images acquired at 1mm isotropic resolution using a three-dimensional (3D) magnetisation-prepared rapid-acquisition gradient-echo (MPRAGE) acquisition (Miller et al., 2016). T1-based image-derived phenotypes (IDPs) were generated for the volumes of major tissue types of the whole brain and for specific structures. This study utilised the cortical brain volume IDPs.

Initially, we combined smaller cortical brain regions to make 23 regions, similar to Harris et al. (2022). Where there was a mismatch between the cortical regions described in Harris et al. (2022) and those in UK Biobank, we grouped brain regions so that they matched Harris’ method as closely as was possible (please see Supplementary Materials for a full list of regional groupings). Next, data from each hemisphere was combined to make a single region, for example the left and right superior frontal gyrus were combined to make a total superior frontal gyrus variable. This decision was taken as depression affects the brain in both hemispheres similarly. Finally, if regions were split further, for example with anterior or posterior divisions, these were also combined to form a single region, for example the left and right anterior supramarginal gyri were combined to make a total anterior supramarginal gyri variable, the left and right posterior supramarginal gyri were combined to make a total posterior supramarginal gyri variable and then the anterior and posterior variables were combined to make a total supramarginal gyrus variable.

RDS-4 questions were extracted from UK Biobank online follow-up mental health questions (UK Biobank codes 2050, 2060, 2070, 2080). The questions are:

1. Frequency of depressed mood in last 2 weeks
2. Frequency of unenthusiasm/disinterest in last 2 weeks
3. Frequency of tenseness/restlessness in last 2 weeks
4. Frequency of tiredness/lethargy in last 2 weeks.

The four RDS-4 questions were combined to give an overall RDS score. Each question used the following scoring:

1. Not at all
2. Several days
3. More than half the days
4. Nearly every day

Responses were scored one point for “Not at all” through to four points for “Nearly every day” and were summed to give an overall RDS-4 score between 4 and 16.

Responses “do not know” (-1) and “prefer not to answer” (-3) for each of the RDS-4 questions were coded as missing data. Missing data was coded in the same way for education, income, and ethnicity. Ethnicity codes 1001 (white British), 1002 (white Irish) and 1003 (any other white background) were combined to create the “White” variable, other ethnicities were combined in the same way, and then coded as a binary variable with 1 as “White” and 0 as “Not white”.

Sex is defined in the UK Biobank as biological sex and is split into male or female. This does not account for gender identity.

SES was measured using education and income. Education is defined as the highest educational attainment in which individuals were categorised as either “College or higher” or “Not higher education” (reference category). Income refers to the average total household income before tax. This variable was split up into five categories: Less than 18,000; 18,000 to 30,999; 31,000 to 51,999; 52,000 to 100,000; and Greater than 100,000.

Multivariate regression analyses were conducted in StataIC 14 to test the association between depression and regional cortical volume in 6 models. Model 1 included age, sex, and ethnicity as covariates, while Model 2 further adjusted for education and income. These analyses were then conducted separately for males and females, with Model 3 and 5 being as Model 1 but for females and males respectively. Models 4 and 6 were as Model 2 but for females and males respectively. These six models were run on each of the 23 brain regions, resulting in 132 models in total. Each model had regional cortical volume as the outcome, and the predictors varied as described above. P-values were Bonferroni-corrected. Multivariate regressions analyses were also conducted to examine the interaction effects between sex and depression for each cortical brain region and then for sex, depression and income for each region.

## Results

We found that higher depression scores were associated with lower cortical volume in eight out of 23 (34.78%) regions when we adjusted the analysis with age and sex in Model 1 (blue dots and lines in Fig. 2). However, this relationship only remained significant in one brain area, the precentral gyrus (b=-43.96, p < .001), when we included education and income in Model 2 (red dots and lines in Fig. 2). Our findings suggest that the relationships between depression scores and the seven brain areas that were no longer significant (the frontal pole; the postcentral gyrus; the supramarginal gyrus; the angular gyrus; the insular cortex; the frontal orbital cortex; and the parahippocampal gyrus) were potentially confounded by education and income.

**Figure 2:**
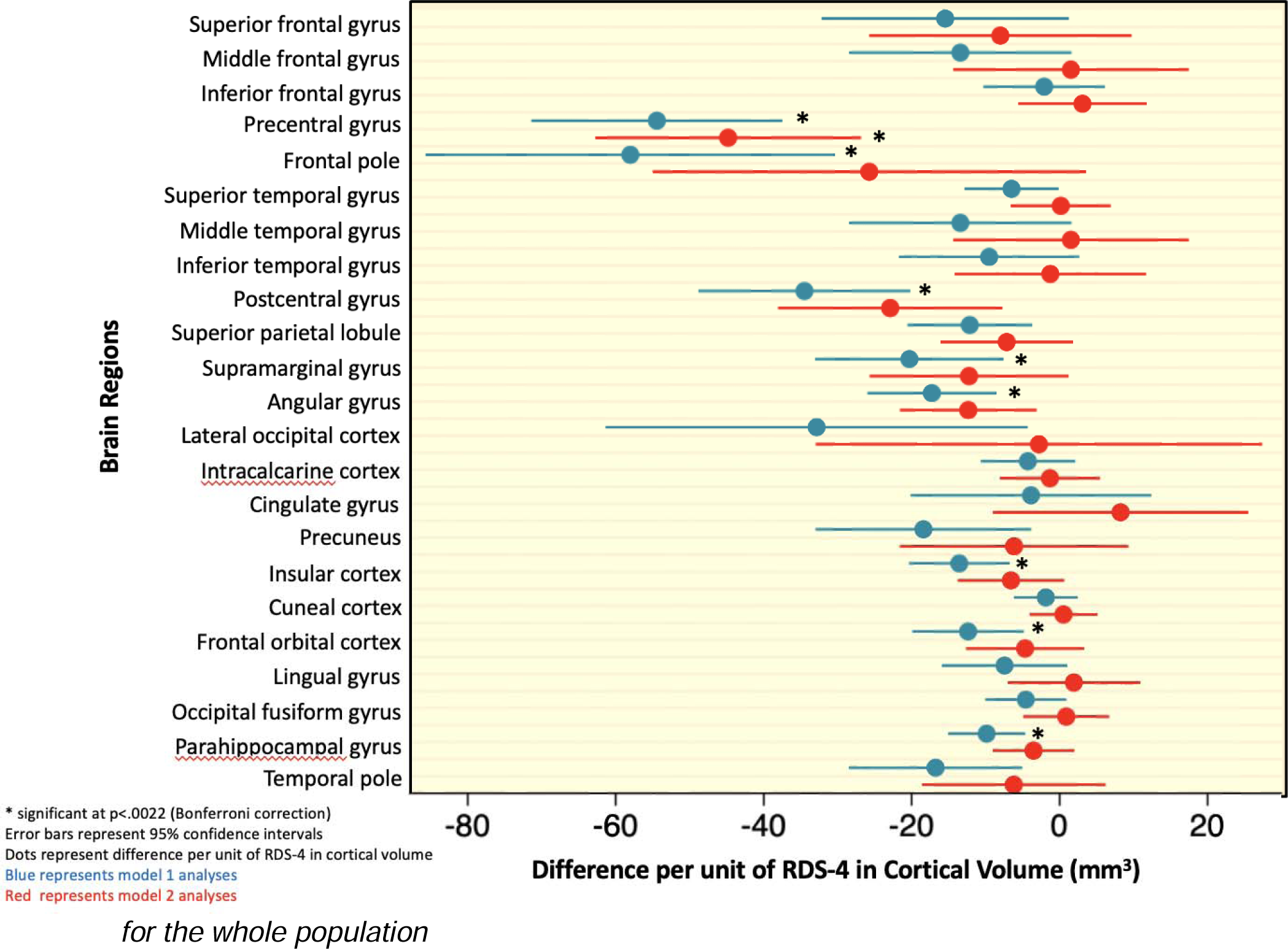
Model comparisons of associations between depression and brain regions for the whole population.

Among females, three regions, the precentral (b=-51.07, p<.001); postcentral (b=-28.92, p=.002); and angular (b=-17.08, p=.002) gyri, had significant negative associations with depression scores when we included only age and sex in the analysis (Model 3, blue dots and lines, Fig. 3). This significant negative relationship was only maintained in the postcentral gyrus (b=-42.43, p<.001) when education and income were included (Model 4, pink dots and lines, Fig. 3).

**Figure 3:**
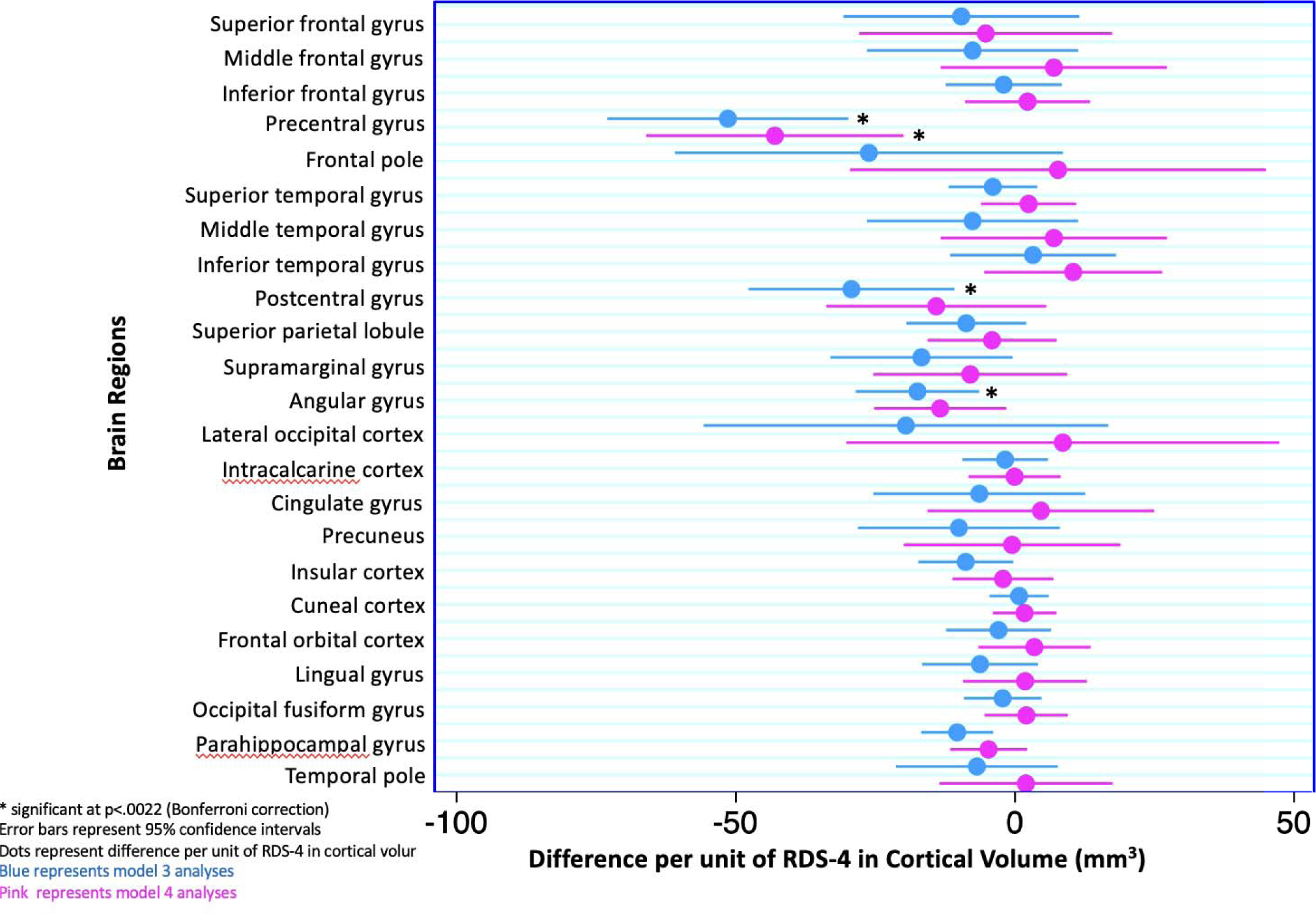
Model comparisons of associations between depression and brain areas for females.

Among males, there were six regions that had significant negative associations with depression scores when only age and sex were included in the analysis (Model 5, blue dots and lines, Fig. 4). However, this relationship became non-significant in four of these regions when education and income were included (Model 6, red dots and lines, Fig.4). The regions that still showed significant negative relationships in Model 6 were the precentral gyrus (b=-46.35, p>.001) and the frontal pole (b=-71.44, p=.002).

**Figure 4:**
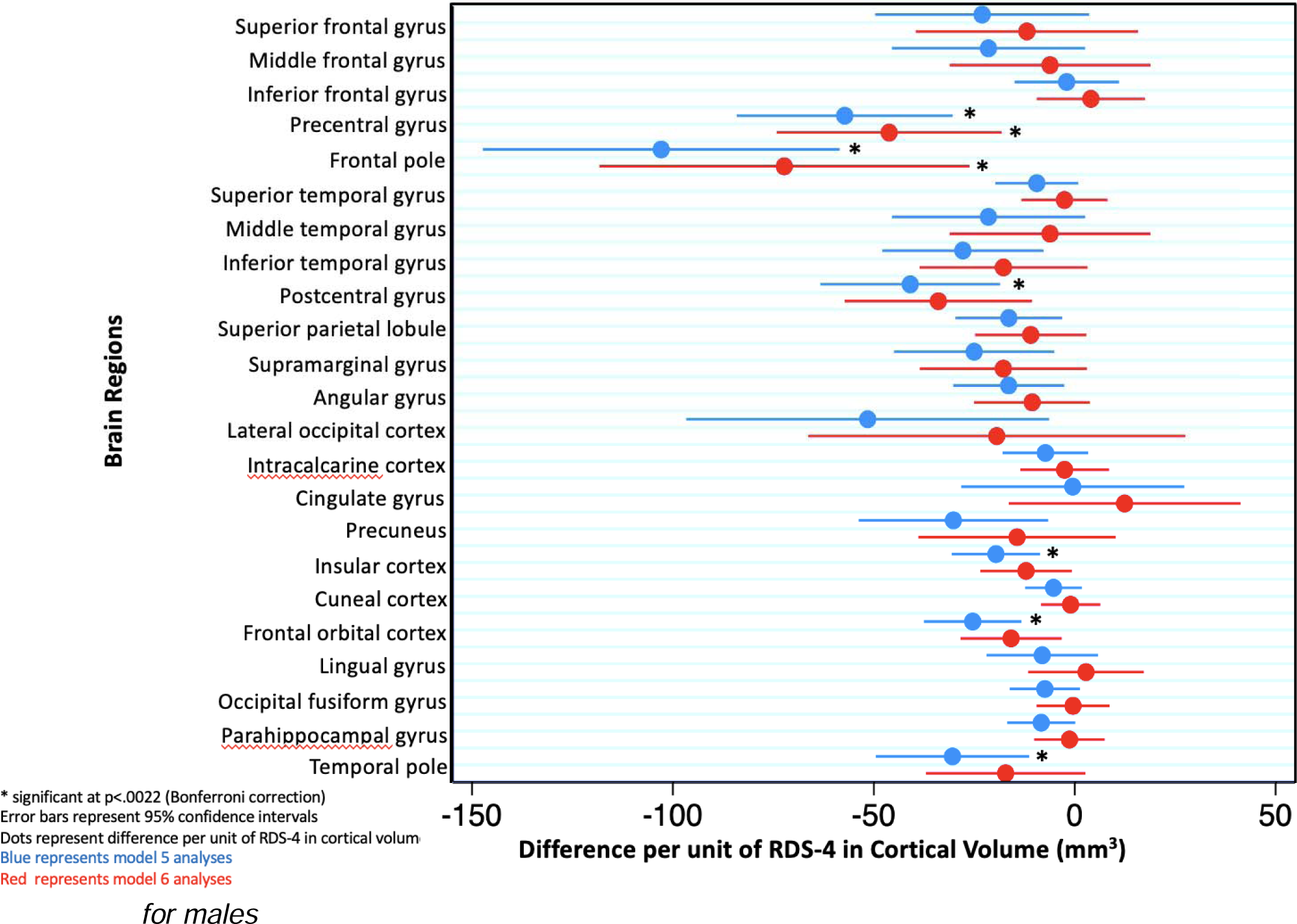
Model comparisons of associations between depression and brain areas for males.

When the interaction terms between sex and depression were added in Model 7, the female by depression scores terms were significant for reduced volume in the inferior temporal gyrus; cuneal cortex; and frontal orbital cortex. These findings suggest significant differences in the relationship between brain volume and depression for males and females in these regions.

## Discussion

We found a significant relationship between cortical volume and depression score in multiple brain regions in a UK Biobank dataset. However, this relationship was affected by the inclusion of education and income and separating the analysis by sex. This is perhaps unsurprising given the fact that socioeconomic factors have been found to impact on the health of males and females differentially (Wardle et al., 2002; Backholer et al., 2017; Park et al., 2012; Gassen et al., 2021) and highlights the fact that social factors need to be considered when examining the relationship between biological factors and mental health.

### Depression and Brain Structure

The precentral gyrus was robustly related to depression score across all analyses and after accounting for SES in both sexes. While this is not one of the regions that are most often associated with depression in the literature, it is in-line with some previous research suggesting a link between this region and depression (Song et al., 2022; Taki et al., 2005; Zhang et al., 2012; Schmaal et al., 2017). The precentral gyrus is the motor centre of the brain and is considered to be the area responsible for voluntary movement. However, it may also be involved in cognitive processing, including working memory (Ren et al., 2019; Yue et al., 2019), implicit learning (Rostami et al., 2009) and silent reading (Kaestner et al., 2021). One study also implicated this region in negative attributional bias (Blackwood et al., 2000), a negative cognitive style that has been implicated in depression (Hu et al., 2015; Moritz et al., 2007; Beck, 2008; Diez-Alegría et al., 2006). The precentral gyrus is also involved in response inhibition, thus Zhang et al. (2012) postulated that the reductions they saw in GMV in the precentral gyrus in individuals with a cognitive vulnerability to depression or diagnosed MDD may demonstrate an abnormality in the ability to inhibit negative attributional bias.

The postcentral gyrus was related to depression score across all analyses that did not include education and income (Models 1, 3 and 5). Again, while this region may not be one of the main regions associated with depression in previous research, there are some studies that have found a relationship. Chen et al. (2016) found that, when compared with age and gender matched healthy controls, drug-naïve individuals who were experiencing their first episode of MDD displayed significantly increased grey matter volume in the right postcentral gyrus. Also, Peng et al. (2019) found that individuals with anxious depression had an increased GMV in the left postcentral gyrus, compared with individuals with non-anxious depression, and that these volumes were positively correlated with severity of depression and anxiety symptoms in individuals with anxious depression. The postcentral gyrus has also been shown to be affected by SES, which may explain why the relationships with depression score in this region were no longer significant when education and income were added into the models. Dufford et al. (2021) found that childhood SES (as measured by income-to-needs ratio) had a positive, prospective relationship with cortical thickness in the postcentral gyrus in adulthood. Mackey et al. (2015) found that students from higher incomes had greater postcentral gyrus cortical volume than those from lower incomes.

Each of the significant relationships found in our analyses were negative, meaning that as depression scores increased, cortical volume in those specific regions decreased, which is consistent with much of the literature (e.g. Wu et al., 2023; McKinnon et al., 2009; Han et al., 2014; Boes et al., 2008; Botteron et al., 2002; Bremner et al., 2002; Lai et al., 2000). There is, however, some research that has found an increase in cortical volume in depression, in regions such as the precuneus, cingulate gyrus, middle frontal gyrus, and angular gyrus (Leung et al., 2009; Scheuerecker et al., 2010; Yuan et al., 2008). These findings are inconsistent with the current study which found a decrease in cortical volume in the precuneus when considering the whole population (Model 1) and males only (Model 5) without including education and income. Our study found no significant relationship between depression scores and volume of the cingulate and middle frontal gyri in any analyses and found a negative relationship between volumes of the angular gyrus and depression scores in the whole population (Model 1) and females individually (Model 3) when SES was not considered. However, the previously mentioned studies (Leung et al., 2009; Scheuerecker et al., 2010; Yuan et al., 2008) differed from ours in that they had much smaller sample sizes (fewer than 20 patients in each study) and used individuals with a diagnosis of MDD. Participants in Leung et al. (2009) and Yuan et al. (2008)’s studies also were all taking anti-depressant medication, whereas we did not have the information regarding psychiatric medication in this study. Indeed, it has been suggested that inconsistency in terms of increases and decreases in cortical volume may be due to such factors as medication effects and chronicity of patients (Qiu et al., 2014). In addition, except for Yuan et al. (2008), the studies mentioned had participants with average ages that were much younger than those in the current study, and age of participants had also been determined to impact reported cortical volume (Qiu et al., 2014).

### Comparison with Harris et al. (2022)

Our results are somewhat consistent with the associations that Harris et al. (2022) found between depression and cortical volume. They also found a relationship between the volumes of the precentral gyrus and self-reported depression. In addition, they found a significant relationship between the orbitofrontal cortex and self-reported depression, although these did not survive false discovery rate (FDR) correction. For the frontal orbital cortex, we found a significant relationship between cortical volume and RDS-4 in Model 1 (both sexes without considering education and income) and Model 5 (males only, without education and income). There was no relationship found for the frontal orbital cortex volume in either of our female models. The volume of the precentral gyrus, on the other hand, had a robust relationship with RDS-4 across all analyses in this study. Interestingly, Harris et al. (2022) found no relationship between the cortical volume of postcentral gyrus and any depression measure, whereas we found evidence of a relationship between postcentral gyrus volume and RDS-4 for each of our models that did not include SES (Models 1, 3 and 5). They found an association between the cortical volume of the middle temporal gyrus and self-reported depression, which did survive correction, whereas we didn’t find any associations with the volume of the middle temporal gyrus and RDS-4 in any model. None of their effects of cortical volume with any phenotype of depression, other than self-reported depression, survived FDR correction.

The fact that some of our results were consistent with Harris et al., (2022) however some were not, perhaps suggests that the RDS-4 is related to brain structure via a different mechanism than the self-report measures of lifetime depression used in Harris et al. (2022). This is consistent with the fact that Harris et al. (2022) reported that some of the phenotypes of depression showed different relationships with brain structure.

The RDS-4 was chosen as it has been shown to be comparable to the PHQ-9 (Dutt et al., 2022), a well-validated measure of depression. Dutt et al. (2022) reported a “stable and approximately linear mapping” between the two. The RDS-4 was also available for nearly all UK Biobank participants, whereas the PHQ-9 was only available for approximately 30% of participants (Amin et al., 2023). The PHQ-9 was also taken independently of participants’ brain scans, with a time discrepancy that was highly inconsistent across subjects, which means that it is not the most suitable measure of state depression in this study (Dutt et al., 2022). Interestingly, when the PHQ and RDS-4 were obtained concurrently, the correlation is high (0.9), however in the UK Biobank, due to the gap in acquisition times, there is a much lower correlation (0.6) (Dutt et al., 2022). Again, emphasising the need measures to be obtained concurrently.

### The effect of Education and Income

Education and income were chosen as the SES variables as they have been used in multiple papers using the same dataset and looking at the effect of SES on health (Shen et al., 2018; Petermann-Rocha et al., 2020; Dong et al., 2022. They have also both been associated with depression in previous studies (Rai et al., 2013; Akhtar-Danesh et al., 2007; Romans et al., 2011). It has also been demonstrated that combining education and income into a composite measure of socioeconomic position creates a more robust estimate of the social gradient of health than considering them separately (Lindberg et al., 2022).

There are several regions that were associated with depression in the models that did not account for SES, but this relationship did not survive once these factors were included, suggesting that they are related to or affected by education and/or income in some way. Given that these regions vary quite widely in terms of both their location and function, it is difficult to come to a definitive interpretation of these findings. However, they have been related to both depression and SES in previous literature. (Bludau et al., 2016; Machlin et al., 2020; Suh et al., 2019; McDermott et al., 2019; Kang et al., 2023; Yaple & Yu, 2020; Schnellbächer et al., 2022; Lai et al., 2000; Holz et al., 2015; Zeng et al., 2012; Jednoróg et al., 2012; Liang et al., 2020; Loued-Khenissi et al., 2022).

Socioeconomic factors have also been shown to affect whole brain health and development, so it stands to reason that there would be a variety of areas affected. For example, parental SES has been shown to affect in vivo fetal neurodevelopment (Lu et al., 2021). Low SES has also been suggested to be characterised by lower brain volume and slower rates of change throughout brain maturation (Rakesh et al., 2023). The fact that SES in this study affected the relationship between brain structure and depression is in line with previous research suggesting a relationship between both brain structure and social factors and depression and social factors, as previously discussed, and emphasises the need to consider all three factors when researching these topics.

### Limitations

The current study did not exclude participants diagnosed with other conditions that could affect the brain. However, given the large, healthy sample, this hopefully will not have substantially affected the results. We also did not control for anxiety, which is strongly comorbid with depression, with 40-50% of individuals with MDD also suffering with anxiety (Xia et al., 2018) and the presence of comorbid anxiety has been shown to alter the relationship between depression and brain structure (Espinoza Oyarce et al., 2020). It also must be noted that the UK Biobank is limited in its diversity in terms of ethnicity, age and income and does not represent the general population on several sociodemographic, physical, lifestyle and health-related factors, with evidence of a healthy-volunteer selection bias (Fry et al., 2017). However, this is common in many imaging studies and the large sample size and heterogeneity of measures allow for generalisations to other populations without the sample being representative of the whole population (Fry et al., 2017). Since the UK Biobank was not designed specifically to investigate depression, many covariates were not available that may have been useful to consider, such as details of therapeutic or pharmacological treatments. Although the RDS-4 has been validated against commonly used depression scales such as the PHQ-9, it is a short-form questionnaire. Future work involving more comprehensive measures of depression would add to the literature. In terms of SES, there are other measures within the UK Biobank such as area deprivation, occupation, job class and employment status that we did not consider, that may have yielded different results. There is also research suggesting that education and income are differentially related to mental health depending on the culture being examined. Araya et al. (2003) examined the relationship between socioeconomic measures and common mental disorders and found a strong, negative independent association between education and common mental disorders but found no association between income and common mental disorders when adjusting for other socioeconomic variables. This contrasts with some British studies that have found income, but not education to be associated with common mental disorders (e.g. Davey Smith et al., 1998; Muller et al., 2002), suggesting that measures might represent different concepts across different cultures. In future research it would be beneficial to look at education and income separately and see if one or the other is contributing to the effects seen more.

### Future Work

The next step is to further consider how these socioeconomic factors combine with structural correlates to predict depression and to gain a better understanding of the relative contribution of socioeconomic factors and brain structure to depression. This study has examined current depressive symptoms and their relationship to cortical volume. Further research is needed to determine whether this relationship is transient or stable when depressive symptoms change. Future research should involve large-scale, longitudinal cohort studies with rich neuroimaging, psychological and sociodemographic measures to fully elucidate the individual contributions of each of the variables in these relationships. The question remains as to whether education and income are causally related to brain structure, or whether they are acting as proxy measures for other factors such as nutrition, Body Mass Index (BMI) or IQ.

## Conclusion

This study is, to our knowledge, the first study to examine the relationship between depression, cortical volume, and SES in a middle-aged to older sample of this size. Given that SES appears to impact the relationship between depression and brain structure, this may have implications for interventions aimed at combatting depression. It could be that certain treatments are less effective for individuals from certain socioeconomic backgrounds or that targeting poverty, or deprivation has a substantial impact on treatment outcomes, beyond traditional pharmacological or therapeutic interventions. Furthermore, we posit that SES should be a consideration for clinicians and scientists working with imaging data in these populations.

## Data Availability

All data is the property of UK Biobank and is available via application

