## Supplemental Table for "Depression, Brain Structure and Socioeconomic Status: A UK Biobank Study"

**Supplementary Materials**

A table to show the combinations of smaller cortical regions to make the final 23 cortical regions used in our analyses

| **Cortical Region** | **Combined regions** |
| --- | --- |
| Superior frontal gyrus | Left + right superior frontal gyrus |
| Middle frontal gyrus | Left + right middle frontal gyrus |
| Inferior frontal gyrus | Left + right inferior frontal gyrus pars triangularis + left + right inferior frontal gyrus pars opercularis |
| Precentral gyrus | Left + right precentral gyrus |
| Frontal pole | Left + right frontal pole |
| Superior temporal gyrus | Left + right superior temporal gyrus anterior division + left + right superior temporal gyrus posterior division |
| Middle temporal gyrus | Left + right middle temporal gyrus anterior division + left + right middle temporal gyrus posterior division + left + right middle temporal gyrus temporooccipital part |
| Inferior temporal gyrus | Left + right inferior temporal gyrus anterior division + left and right inferior temporal gyrus posterior division + left + right inferior temporal gyrus temporooccipital part |
| Postcentral gyrus | Left + right postcentral gyrus |
| Superior parietal cortex (named superior parietal lobule in UK Biobank) | Left + right superior parietal lobule |
| Supramarginal gyrus | Left + right supramarginal gyrus anterior division + left + right supramarginal gyrus posterior division |
| Angular gyrus | Left + right angular gyrus |
| Lateral occipital cortex | Left + right lateral occipital cortex superior division + left + right lateral occipital cortex inferior division |
| Intracalcarine cortex | Left + right intracalcarine cortex |
| Cingulate gyrus | Left + right cingulate gyrus anterior division + left + right cingulate gyrus posterior division |
| Precuneus (precuneous cortex in UK Biobank) | Left + right precuneous cortex |
| Insula (insular cortex in UK Biobank) | Left + right insular cortex |
| Cuneus (cuneal cortex in UK Biobank) | Left + right cuneal cortex |
| Orbitofrontal cortex (frontal orbital cortex in UK Biobank) | Left + right frontal orbital cortex |
| Lingual cortex (lingual gyrus in UK Biobank) | Left + right lingual gyrus |
| Fusiform gyrus (occipital fusiform gyrus in UK Biobank) | Left + right occipital fusiform gyrus |
| Parahippocampal gyrus | Left + right parahippocampal gyrus anterior division + left + right parahippocampal gyrus posterior division |
| Temporal pole | Left + right temporal pole |
